# Lassa fever epidemiology and predictors of mortality in Ebonyi State, Nigeria – A five-year retrospective analysis from 2019 to 2023

**DOI:** 10.64898/2026.07.02.26357189

**Authors:** Eric C. Nwojiji, Ogbonna N. Nwambeke, Pin-Wei Shih, Clement U. Chukwunenye, Elizabeth C. Odeh, Moses I. Ekuma, Benedict N. Azuogu, Michael O. Iroezindu, Laurens Liesenborghs, Christophe Van Dijck

**Affiliations:** Ebonyi State Ministry of Health Abakaliki, Nigeria; Department of Community Medicine, Faculty of Clinical Medicine, College of Medicine, Alex Ekwueme Federal University Ndufu-Alike, Ebonyi State, Nigeria; Institute of Tropical Medicine, Antwerp; Department of Public Health, Institute of Tropical Medicine Antwerp, Belgium; Department of clinical sciences, Institute of Tropical Medicine Antwerp, Belgium; Department of Epidemiology, Medécins Sans Frontiére Belgium; Alex Ekwueme Federal University Teaching Hospital Abakaliki, Nigeria; Virology laboratory, Alex Ekwueme Federal University Teaching Hospital Abakaliki, Nigeria; Department of Community Health, Alex Ekwueme Federal University Teaching Hospital Abakaliki, Nigeria; David Umahi Federal University of Health Sciences, Uburu Nigeria; School of Public Health, Faculty of Community and Health Sciences, University of the Western Cape, Bellville, South Africa; Virology, Antiviral drug & Vaccine research group, Department of Microbiology, Immunology and Transplantation, KU Leuven, Leuven, Belgium

**Keywords:** Lassa fever, Trend, Spatial distribution, Time series, Ebonyi State

## Abstract

Lassa fever (LF) is a viral disease that is widespread throughout West Africa, with significant global health consequences. Nigeria bears the highest burden (1309 confirmed cases in 2024) of LF among all endemic countries. Within Nigeria, Ebonyi State bears a high burden of cases. Clinical observations suggest that there may be an increase in the geographic spread and case fatality rate of LF across the State. Ebonyi State case-based data on confirmed cases from 2019 to 2023 collated on the Nigeria Centre for Disease Control and Prevention (NCDC) Surveillance, Outbreak Response Management and Analysis System (SORMAS) platform were analyzed. Descriptive statistics, spatial distribution and time series analysis were performed. Multivariate logistic regression analysis was used to identify predictors of LF mortality and factors associated with PCR positivity. A total of 1,624 suspected cases were reported, 1,343 and 273 were laboratory negative and positive respectively, while 8 probable cases were reported. The yearly number of cases remained stable throughout the study period. Out of the 273 confirmed, 107 died from LF, resulting in a case fatality rate (CFR) of 39.2%. CFR increased non-significantly over time, ranging from 28.6% to 55.8%. Variations in geographic distribution were observed; in 2019 ten local government areas (LGAs) were affected compared to twelve in 2020. A higher incidence was observed between January and March annually. Age above 44 years, bleeding and seizures were significant predictors of mortality. Lower incidence of cases was consistently reported in the Southern LGAs. PCR positivity was associated with individuals who reside in Ebonyi LGA and who have had contact with confirmed cases. The increase in CFR and identification of high-burden areas will help shape policies, allocate resources and provide actionable intervention strategies to combat LF in the State.

## Introduction

Lassa fever (LF) is a viral disease that is widespread throughout West Africa, and it has significant global health consequences. (1,2). The causal agent is the Lassa virus (LASV), a single-stranded ribonucleic acid (RNA) virus from the *Arenaviridae* family. The multimammate rat of the genus *Mastomys* is the main reservoir of LASV, but the virus has also been found in other rodents (3,4) and there is some serological evidence of LASV infection in non-rodent species (5). Seroprevalence studies have estimated that LASV is responsible for 100,000-300,000 cases annually and 5,000 deaths (6) in a region where Lassa fever is endemic. However, it is unclear how far these projections are correct. LF therefore imposes a significant burden in endemic regions, accounting for 0.7% of hospital admissions and 25% of maternal deaths during peak seasons (7–10). Outbreaks occur very often, with a peak occurring during the dry season months of November to April (11)

Human LASV infection occurs when manipulating infected materials or when consuming contaminated food and infected rodent meat. Person-to-person transmission can occur from direct contact with blood, urine, excretion, or tissue of infected subjects (12,13). Healthcare workers are at high risk of exposure; therefore, protective equipment should be used for sample handling or when caring for infected patients. Airborne transmission has been suggested to play a role (14), although this remains controversial.

Diagnosis in Ebonyi State today, is primarily conducted through real-time reverse transcriptase polymerase chain reaction (RT-PCR) (15) using Altona 2.0 reagents in a magnetic induction cycler (MIC). Ribavirin and general supportive therapy may improve outcomes, especially if instituted early (16).

Lassa fever surveillance in Ebonyi State and Nigeria at large is done through the Integrated Disease Surveillance and Response (IDSR) system adopted in September 1998 with the goal to improve the ability of health facilities and public health authorities at Local Government Area (LGA), State and national levels to detect and respond to diseases and conditions that cause high level of illness, disability and death. The Surveillance Outbreak Response Management & Analysis System (SORMAS), adapted as the electronic IDSR, established in 2014/15 following outbreak of Ebola virus disease in West Africa was expanded to include priority diseases such as LF due to its high burden in the country. Ebonyi State LF reporting flows from the community to the national level (Nigeria Centre for Disease Control and Prevention, Federal Ministry of Health) through the health facility, LGA Disease Surveillance and Notification Officers (DSNOs), and State levels (State DSNO, State epidemiologist) (17) using standard case definitions. After the implementation of SORMAS in Nigeria, a significant increase in the number of LF cases was reported in 23 out of 36 States (8). In 2021, a study indicated that LF is now endemic in 32 out of 36 States of Nigeria (18).

Clinical observations suggest that there may be an increase in the geographic spread and case fatality rate of LF across Ebonyi State. However, due to limited evidence, this study aims to analyze and describe the temporal evolution in the number of reported LF cases in Ebonyi State, their geographic distribution, and associated CFR. This study also assesses the factors associated with PCR positivity and predictors of LF-associated mortality in Ebonyi State from 2019 to 2023. The findings of this study will help relevant authorities develop appropriate strategic policies and targeted interventions to combat the infection.

## Methods

### Study area

Ebonyi State is one of the 36 States in Nigeria, located in the south-east of the country. She has thirteen LGAs and an estimated population of 2.5 million (22,23). Approximately 80% of its inhabitants are subsistence farmers and are rural dwellers. (19).

### Study design & data source

This is a retrospective study of surveillance data on LF cases in Ebonyi State employing a descriptive and analytical approach. The data from January 2019 to December 2023 was extracted from the NCDC-SORMAS database and provided by the Ebonyi State Ministry of Health in an excel format for analysis. The reporting of LF cases in Ebonyi State is based on case definitions a**s** defined by the NCDC (Table 1).

**Table 1.** Lassa fever case definitions.

|  | Definitions |
| --- | --- |
| Suspect | A suspected case defined as a patient with fever for 3-21 days with a measured temperature of 38°C or more with one or more of the following: vomiting, diarrhea, sore throat, myalgia (muscle pain), generalized body weakness, abnormal bleeding, abdominal pain **OR** in **neonates**: maternal Lassa fever +/- signs and symptoms. |
| Probable | A probable case is considered a suspected case who has one or more of the following complications: hearing loss, facial or neck swelling, seizures, restlessness, confusion, hypotension (SBP< 90mmHg in adults and <70mmHg in |
| | children), oliguria ($<0.5\text{ml/kg/h}$ for 6 hours), abnormal bleeding and any of the following supporting **laboratory features**: proteinuria and/or microscopic hematuria, elevated urea $\geq 45\text{ mg/dl}$ or creatinine $\geq 2\text{ mg}$, elevated transaminases (liver enzymes, ALT & AST), reduced platelets count $\leq 90,000\text{ cells/ml}^3$. |
| Confirmed | A confirmed case is a suspected or probable case with a positive real-time polymerase chain reaction (PCR) test. |

*SBP: systolic blood pressure, ALT: alanine aminotransferase, AST: aspartate aminotransferase*

### Data analysis

The data was summarized using frequencies and proportions for categorical data, and medians with interquartile ranges for continuous variables. A spatial analysis was performed to illustrate the geographic spread of LF and CFR over time. Moran’s I statistic was used to demonstrate geographic clustering of confirmed cases. A time series analysis was employed to describe the seasonality of the infection.

Risk factors associated with LASV PCR positivity among suspected cases were analyzed using logistic regression models. Predictors of LF mortality among confirmed cases were analyzed using logistic regression models. Univariate analyses were conducted for variable selection. The variables with a p-value < 0.2 were kept for inclusion in the multivariate models to ensure that important variables were not omitted. Multivariate logistic regression models were built using the forward approach, adding one variable at a time while evaluating the model’s Akaike information criterion (AIC) after each addition. Data analyses were executed using R version 4.5.0.

### Use of artificial intelligence tools

Some phrases were reconstructed using Grammarly and copilot with prompts like “Please rephrase the following sentence…” Meta AI was used for code troubleshooting and validated using R.

### Ethical approval

Ethics approval was obtained from the Institutional Review Board of the Institute of Tropical Medicine Antwerp, Belgium **(1749/24)** and the Ebonyi State Ministry of Health **(EBSHREC/0061/052)**. Data access was finalized on April 10, 2024. To ensure participant confidentiality, all data were anonymized, and the research team had no access to personal identifiers. Data protection and confidentiality were maintained throughout the study period.

## Results

### Characteristics of reported cases

A total of 1624 suspected cases were reported from 2019 to 2023. Within this period, 273/1624 (16.8%) were PCR-confirmed, while 1343/1624 (82.7%) tested negative on RT-PCR. Eight probable cases were PCR-negative but had clinical symptoms suggestive of a VHF. The highest proportion of confirmed cases was recorded in individuals aged 15-44 years 174/273 (65.6%).

The proportion of males was 138/273 (50.5%), females 133/273 (48.7%), and secondary school students make up 104/273 (38.1%). Aside from students or children with barely any reported occupation, those who reported their occupation as business or trading and manual laborers represented the bulk of the cases 96/273 (35.1%). The highest proportion of confirmed cases was recorded in Abakaliki LGA, 144/273 (52.7%), followed by Izzi 29/273 (10.6%), Ezza North 28/273 (10.3%), and Ikwo 22/273 (8.1%) LGAs (Table 2)..

**Table 2.** Characteristics of reported Lassa fever cases in Ebonyi State from 2019 to 2023.

|  | **Confirmed case**<br>(N=273) | **Probable case**<br>(N=8) | **Negative case**<br>(N=1343) | **Overall**<br>(N=1624) |
| --- | --- | --- | --- | --- |
| **Sex** |  |  |  |  |
| Female | 133 (48.7%) | 4 (50.0%) | 657 (48.9%) | 794 (48.9%) |
| Male | 138 (50.5%) | 4 (50.0%) | 669 (49.8%) | 811 (49.9%) |
| Missing | 2 (0.7%) | 0 (0%) | 17 (1.3%) | 19 (1.2%) |
| **Age Group (years)** |  |  |  |  |
| 0-4 | 12 (4.4%) | 1 (12.5%) | 137 (10.2%) | 150 (9.2%) |
| 5-14 | 27 (9.9%) | 0 (0%) | 173 (12.9%) | 200 (12.3%) |
| 15-24 | 47 (17.2%) | 1 (12.5%) | 266 (19.8%) | 314 (19.3%) |
| 25-34 | 66 (24.2%) | 4 (50.0%) | 276 (20.6%) | 346 (21.3%) |
| 35-44 | 66 (24.2%) | 0 (0%) | 215 (16.0%) | 281 (17.3%) |
| 45-49 | 17 (6.2%) | 1 (12.5%) | 71 (5.3%) | 89 (5.5%) |
| 50+ | 35 (12.8%) | 1 (12.5%) | 175 (13.0%) | 211 (13.0%) |
| Missing | 3 (1.1%) | 0 (0%) | 30 (2.2%) | 33 (2.0%) |
| **Education** |  |  |  |  |
| No Education/In Nursery | 18 (6.6%) | 1 (12.5%) | 132 (9.8%) | 151 (9.3%) |
| Primary | 33 (12.1%) | 0 (0%) | 78 (5.8%) | 111 (6.8%) |
| Secondary | 104 (38.1%) | 2 (25.0%) | 382 (28.4%) | 488 (30.0%) |
| Tertiary | 55 (20.1%) | 0 (0%) | 405 (30.2%) | 460 (28.3%) |
| Missing | 63 (23.1%) | 5 (62.5%) | 346 (25.8%) | 414 (25.5%) |
| **Occupation** |  |  |  |  |
| Business/Trading | 49 (17.9%) | 0 (0%) | 248 (18.5%) | 297 (18.3%) |
| Healthcare | 14 (5.1%) | 0 (0%) | 13 (1.0%) | 27 (1.7%) |
| Housewife | 18 (6.6%) | 1 (12.5%) | 30 (2.2%) | 49 (3.0%) |
| Manual Labor | 47 (17.2%) | 2 (25.0%) | 130 (9.7%) | 179 (11.0%) |
| Students/Children | 83 (30.4%) | 0 (0%) | 512 (38.1%) | 595 (36.6%) |
| Other | 41 (15.0%) | 0 (0%) | 150 (11.2%) | 191 (11.8%) |
| Missing | 21 (7.7%) | 5 (62.5%) | 260 (19.4%) | 286 (17.6%) |
| **Local Government Areas** |  |  |  |  |
| Abakaliki | 144 (52.7%) | 3 (37.5%) | 532 (39.6%) | 679 (41.8%) |
| Afikpo North | 5 (1.8%) | 0 (0%) | 40 (3.0%) | 45 (2.8%) |
| Afikpo South | 1 (0.4%) | 0 (0%) | 20 (1.5%) | 21 (1.3%) |
| Ebonyi | 14 (5.1%) | 0 (0%) | 125 (9.3%) | 139 (8.6%) |
| Ezza North | 28 (10.3%) | 0 (0%) | 67 (5.0%) | 95 (5.8%) |
| Ezza South | 14 (5.1%) | 2 (25.0%) | 48 (3.6%) | 64 (3.9%) |
| Ikwo | 22 (8.1%) | 0 (0%) | 75 (5.6%) | 97 (6.0%) |
| Ishielu | 3 (1.1%) | 0 (0%) | 67 (5.0%) | 70 (4.3%) |
| Ivo | 0 (0%) | 0 (0%) | 7 (0.5%) | 7 (0.4%) |
| Izzi | 29 (10.6%) | 3 (37.5%) | 153 (11.4%) | 185 (11.4%) |
| Ohaozara | 5 (1.8%) | 0 (0%) | 48 (3.6%) | 53 (3.3%) |
| Ohaukwu | 4 (1.5%) | 0 (0%) | 124 (9.2%) | 128 (7.9%) |
| Onicha | 4 (1.5%) | 0 (0%) | 37 (2.8%) | 41 (2.5%) |
| **Year of Investigation** |  |  |  |  |
| 2019 | 55 (20.1%) | 2 (25.0%) | 269 (20.0%) | 326 (20.1%) |
| 2020 | 87 (31.9%) | 4 (50.0%) | 367 (27.3%) | 458 (28.2%) |
| 2021 | 22 (8.1%) | 0 (0%) | 146 (10.9%) | 168 (10.3%) |
| 2022 | 51 (18.7%) | 0 (0%) | 283 (21.1%) | 334 (20.6%) |
| 2023 | 55 (20.1%) | 0 (0%) | 242 (18.0%) | 297 (18.3%) |
| Missing | 3 (1.1%) | 2 (25.0%) | 36 (2.7%) | 41 (2.5%) |
| **Case Outcome** |  |  |  |  |
| Deceased | 107 (39.2%) | 8 (100%) | 16 (1.2%) | 131 (8.1%) |
| Recovered | 134 (49.1%) | 0 (0%) | 1326 (98.7%) | 1460 (89.9%) |
| Missing | 32 (11.7%) | 0 (0%) | 1 (0.1%) | 33 (2.0%) |

PCR – Polymerase chain reaction

### Factors associated with PCR positivity

Comparing suspected cases who tested positive with those that tested negative, we found that individuals with tertiary education were associated with a significantly lower odds of a positive PCR [OR 0.34, 95% CI: 0.13-0.92]. The odds of having a positive PCR were lower in Ebonyi LGA [OR 0.012, 95% CI: 0.15-0.77]. Students and children, when compared to those in business or trading, showed significantly lower odds of being PCR positive [OR 0.53, 95% CI: 0.28-1.00]. Individuals who had contact with an LF case had a higher odds of a positive PCR [OR 2.61, 95% CI: 1.13-5.81].

After adjustments, those residing in Ebonyi LGA and who had contact with a confirmed case were independently associated with PCR positivity (Table 3). Education and occupation variables were excluded from the model due to multicollinearity with the age groups.

**Table 3.**
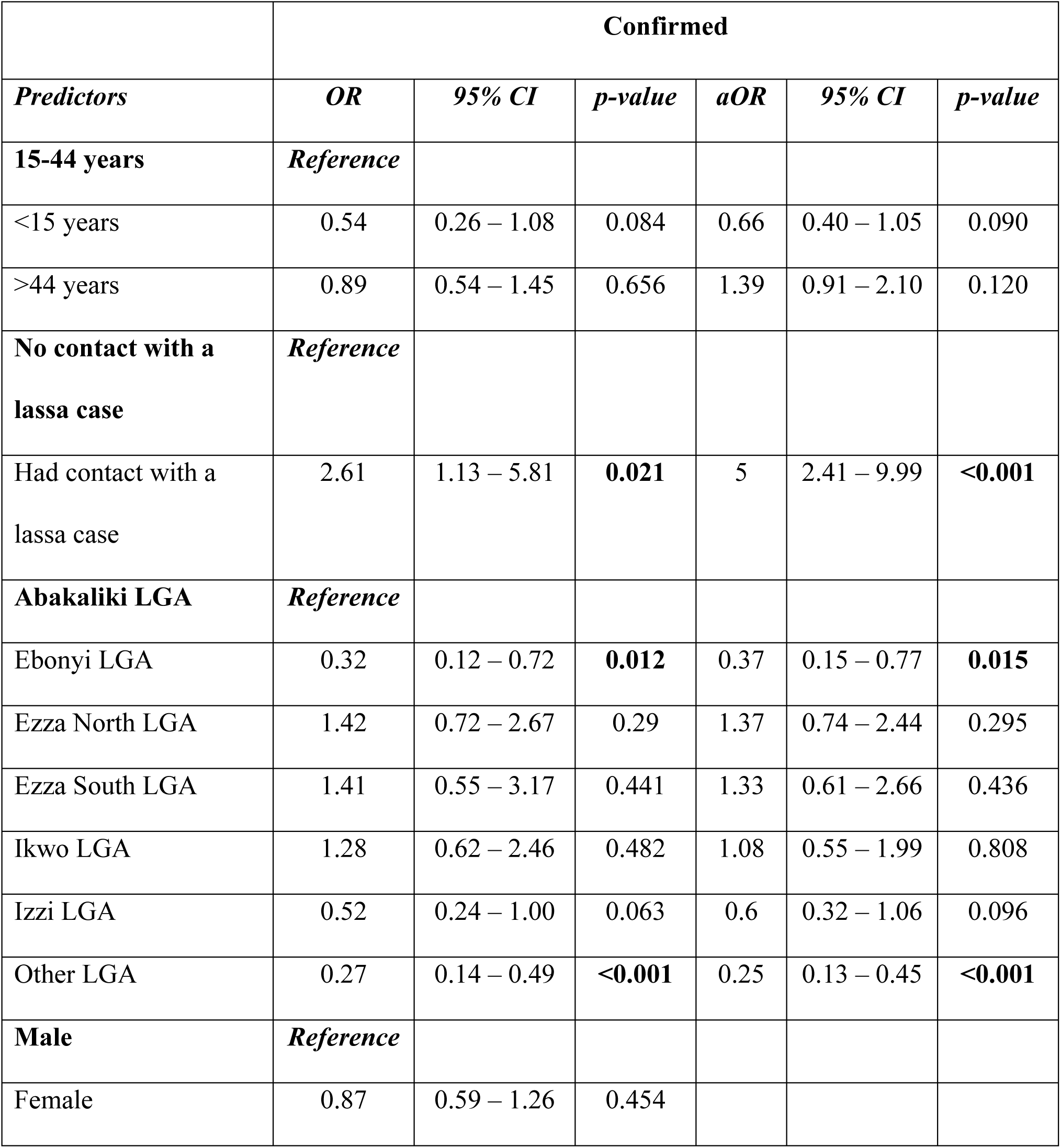
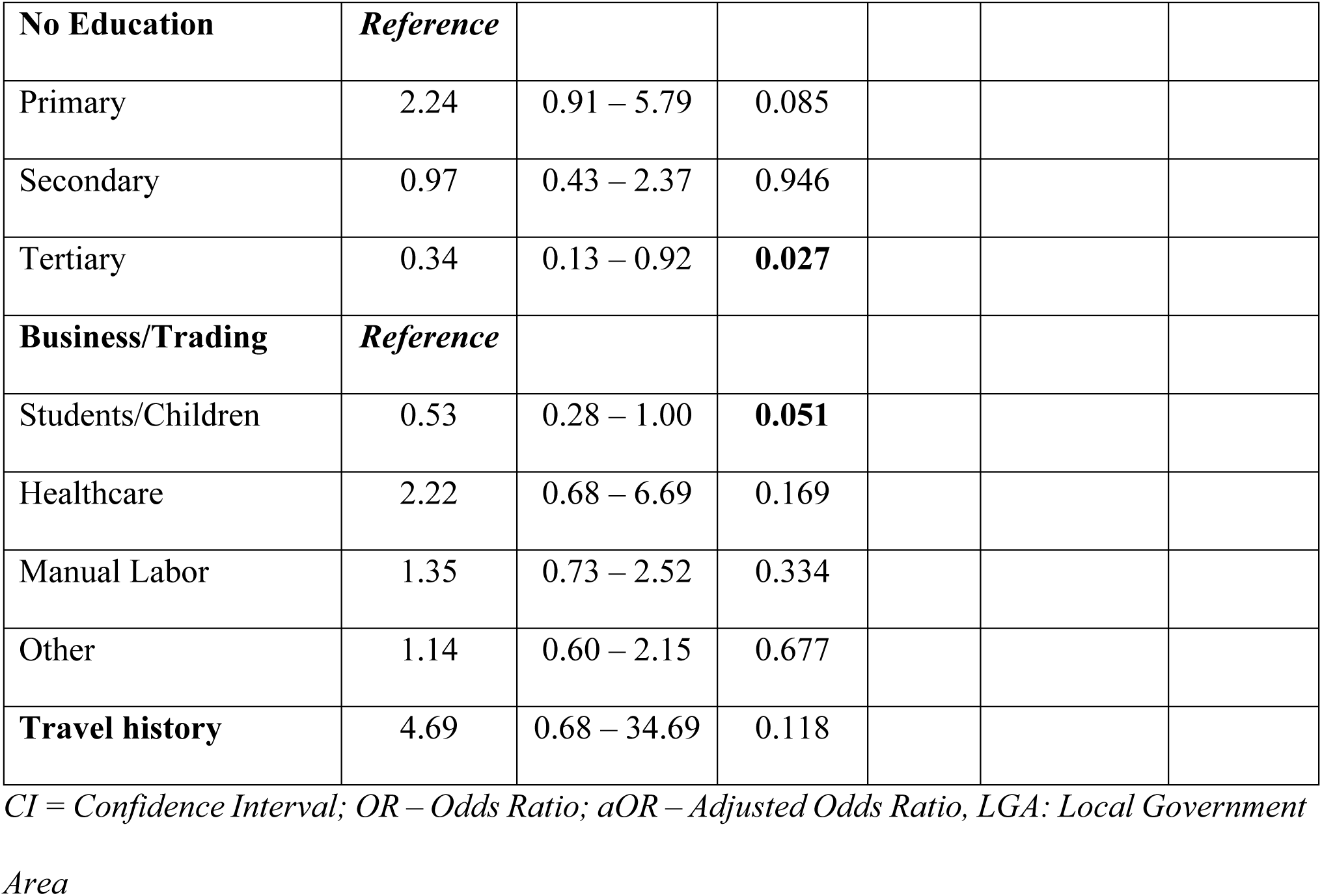
Logistic Regression: Factors associated with Lassa Fever PCR positivity.

### Temporal pattern

Monthly variations in the number of cases per year were observed with seasonal peaks occurring between November and March annually (Fig 1).

**Fig 1.**
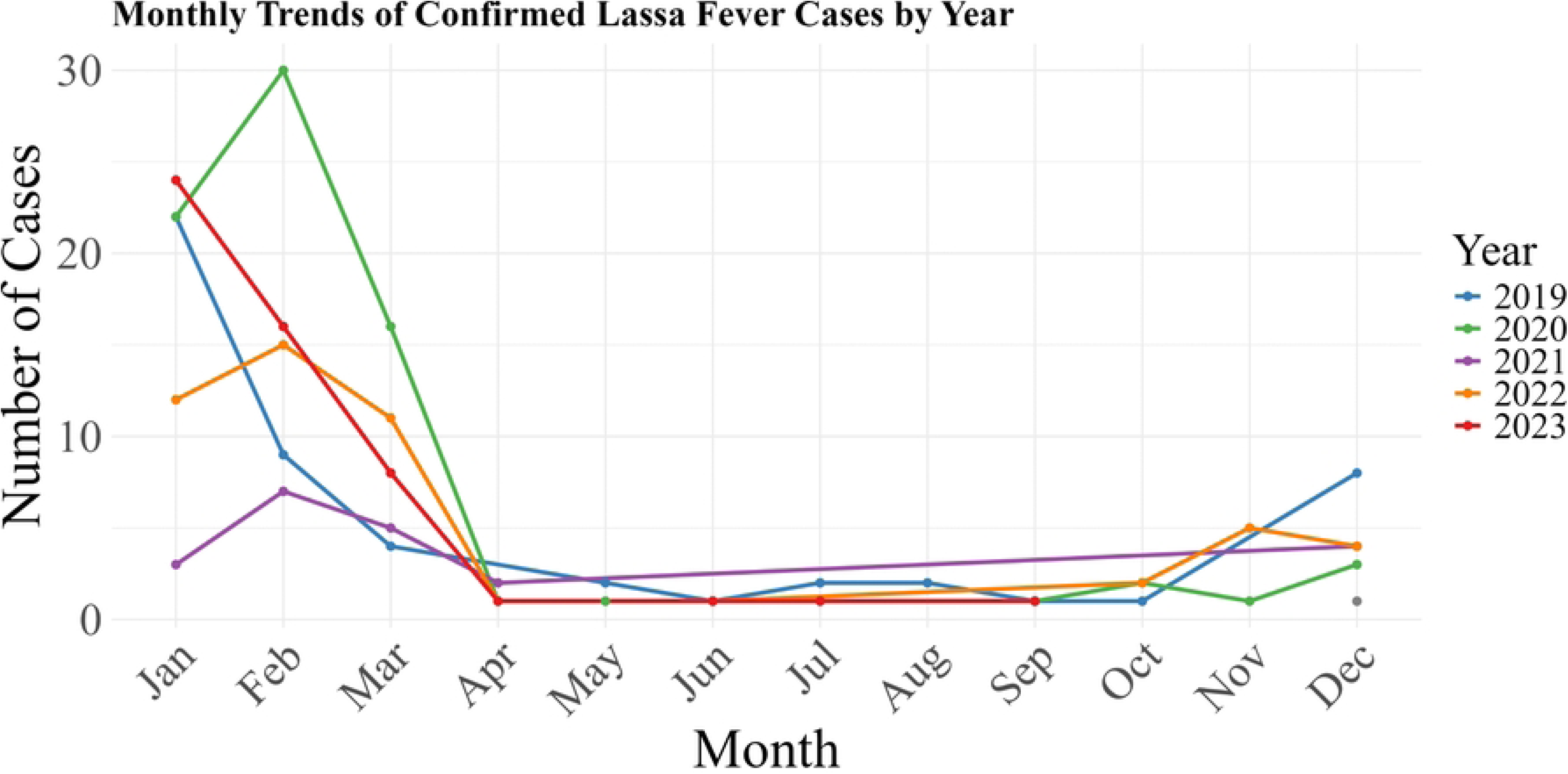
Monthly trend of confirmed Lassa fever cases in Ebonyi State from 2019 to 2023

### Geographic distribution and case fatality rate of confirmed Lassa fever cases

Variation in the geographic distribution of LF across thirteen LGAs was observed, with 10 LGAs affected in 2019 and 11 out of 13 LGAs affected in 2020. However, in 2021, 2022 and 2023, only 9, 8, and 9 LGAs were affected, respectively. Low incidence of cases in the southern LGAs (Fig 2) was consistently observed. Ivo LGA recorded no case during the year under review. A positive Moran’s I (I = 0.11979, p = 0.048) though indicating a weak clustering, also reveal that LGAs with high numbers of confirmed cases are situated next to LGAs with similar burden.

**Fig 2.**
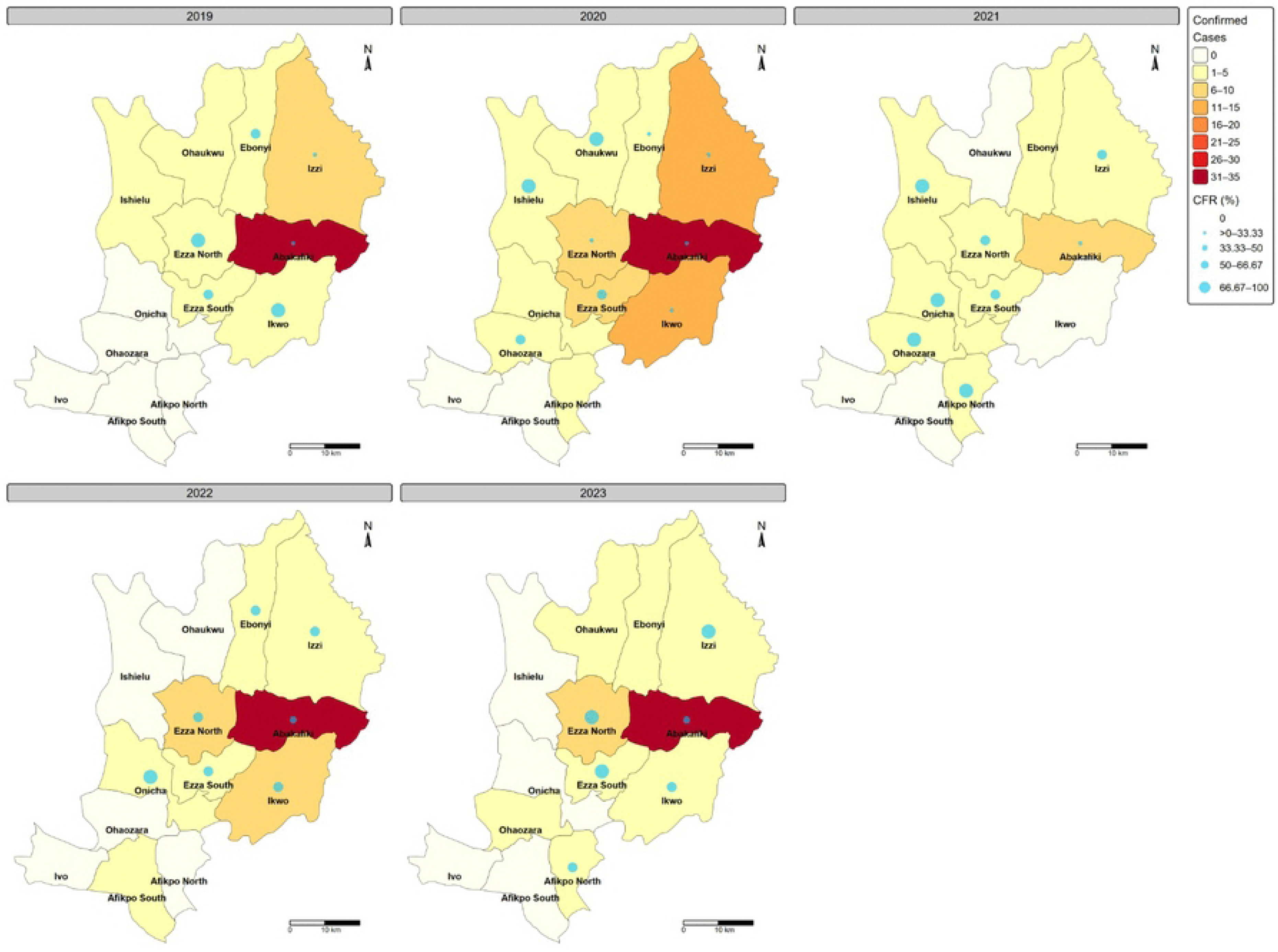
Geographic distribution and case fatality rates of confirmed Lassa fever cases in Ebonyi State from 2019 to 2023.

The CFR for the confirmed cases ranged annually from 28.6% to 55.8% over the five-year period with an overall CFR of 39.2% (107 deaths/273 cases) although 32 observations were unaccounted for.

### Predictors of mortality

Bivariate analysis showed no association between mortality and reporting year (2019–2023). Bleeding [OR 2.27, 95% CI: 1.05–5.00; p*<0.05*], seizure [OR 4.72, 95% CI: 1.63–15.42; p*<0.05*] and age above 44 years [OR 3.00, 95% CI: 1.13–8.29], were all associated with LF mortality (Table 4). Other factors, such as sex, contact with a confirmed case, time to presentation, occupation, and LGA, were not significantly associated with mortality. However, multivariable analysis shows that those aged >44 years [OR 4.12, 95% CI: 1.96–9.10; p*<*0.001] and clinical symptoms such as seizures and bleeding were independently associated with mortality.

**Table 4.** Logistic regression: Predictors of Lassa fever mortality.

| Predictors | Observation (%) | Outcome |  |  |  |  |  |
| --- | --- | --- | --- | --- | --- | --- | --- |
|  |  | OR | 95% CI | p-value | aOR | 95% CI | p-value |
| 15-44 years | 63 (58.8) | Reference |  |  |  |  |  |
| <15 years | 11 (10.2) | 1.94 | 0.54 – 7.0<br>4 | 0.306 | 1.19 | 0.49 – 2.<br>82 | 0.693 |
| >44 years | 32 (29.9) | 3.00 | 1.13 – 8.2<br>9 | **0.029** | 4.12 | 1.96 – 9.<br>10 | **<0.001** |
| **Reporting year** |  | 1.00 | 1.00 – 1.0<br>0 | 0.466 | 1.00 | 1.00 – 1.<br>00 | 0.085 |
| **bleeding** | 43 (40.2) | 2.27 | 1.05 – 5.0<br>0 | **0.038** | 2.12 | 1.14 – 3.<br>99 | **0.018** |
| **Seizure** | 24 (22.4) | 4.72 | 1.63 – 15.<br>42 | **0.006** | 2.59 | 1.13 – 6.<br>24 | **0.028** |
| **shock** | 14 (13.1) | 1.36 | 0.39 – 5.0<br>3 | 0.632 |  |  |  |
| **diarrhea** | 49 (45.8) | 0.80 | 0.38 – 1.6<br>2 | 0.533 |  |  |  |
| **Male** | **55 (51.4)** | **Referenc**<br>**e** |  |  |  |  |  |
| Female | 52 (48.6) | 0.60 | 0.30 – 1.2<br>1 | 0.160 |  |  |  |
| **Business/Tradin**<br>**g** | **24 (22.4)** | **Referenc**<br>**e** |  |  |  |  |  |
| Students/Childre<br>n | 21 (19.6) | 0.46 | 0.15 – 1.3<br>5 | 0.164 |  |  |  |
| Healthcare | 4 (3.7) | 0.79 | 0.15 – 3.8<br>8 | 0.777 |  |  |  |
| Manual Labor | 27 (25.2) | 1.12 | 0.39 – 3.2<br>2 | 0.832 |  |  |  |
| Other |  | 1.53 | 0.51 – 4.6<br>7 | 0.444 |  |  |  |
| **Abakaliki LGA** | **49 (45.8)** | ***Referenc***<br>***e*** |  |  |  |  |  |
| Ebonyi LGA | 4 (3.7) | 0.44 | 0.06 – 2.2<br>4 | 0.356 |  |  |  |
| Ezza North LGA | 16 (15) | 1.95 | 0.67 – 5.8<br>6 | 0.224 |  |  |  |
| Ezza South LGA | 8 (7.5) | 3.57 | 0.75 – 19.<br>37 | 0.118 |  |  |  |
| Ikwo LGA | 11 (10.3) | 3.08 | 0.87 – 11.<br>29 | 0.082 |  |  |  |
| Izzi LGA | 9 (8.4) | 1.02 | 0.22 – 4.2<br>5 | 0.982 |  |  |  |
| Other |  | 1.43 | 0.38 – 5.4<br>6 | 0.596 |  |  |  |
| **No contact with**<br>**a lassa case** | **48 (44.9)** | ***Referenc***<br>***e*** |  |  |  |  |  |
| Had contact with<br>a lassa case | 5 (4.7) | 0.19 | 0.03 – 1.0<br>4 | 0.067 |  |  |  |
| ≤6 days | 7 (6.5) | *Reference* |  |  |  |  |  |
| 7-14 days | 56 (52.3) | 1.55 | 0.47 – 5.6<br>3 | 0.483 |  |  |  |
| 15+ days | 40 (37.4) | 2.14 | 0.63 – 8.1<br>0 | 0.237 |  |  |  |

*CI = Confidence Interval; OR – Odds Ratio; aOR – Adjusted Odds Ratio*

## Discussion

This study describes the demographics, geographic distribution of LF cases in Ebonyi State over time, case fatality rate, predictors of mortality and risk factors associated with PCR positivity. Variation in the geographic distribution of LF across the thirteen LGAs was observed with Ivo LGA recording no case. A non-significantly increasing trend in the fatality rate was noted, and there was no significant association between mortality and time. Fewer cases were consistently observed in the southern LGAs, and there was persistent clustering in Abakaliki LGA with similar burden noted in Ezza North, Izzi, and Ikwo LGAs.

Confirmed cases were not uniformly distributed throughout the years. LF in Ebonyi State exhibited seasonal variability, with peaks observed annually between November and March. Similar findings have been documented (18,20). Ecological factors such as rainfall, bush burning and deforestation activities for farming and meat-hunting purposes during the dry seasons destroy the rodent habitats and encourage their movements into houses, favoring higher cases (21–24). A high incidence of confirmed cases is observed in urban LGAs such as Abakaliki, Izzi, Ikwo and Ezza North, in contrast to the rural LGAs where fewer cases are observed. Persistent clustering in Abakaliki LGA was also noted which may indicate a localized transmission. These findings agree with previous studies (25,26). The urban-rural disparity suggests that urban centres have better access to healthcare, testing capacity and lower fatality rates despite high case numbers.

Being the first to assess the factors associated with PCR positivity in Ebonyi State, this study showed that residing in Ebonyi LGA and having contact with a confirmed case were significantly associated with LF disease. This finding aligns with the LASCOPE cohort study in Nigeria which highlighted that contact with infected individuals particularly households, was a major determinant of disease outcomes (27). Beyond Nigeria, the finding also corresponds with observed patterns in other endemic regions as highlighted by a systematic review of LF in West Africa (28).

An overall CFR of 39.2%, consistent with earlier findings (25) was observed during the five years under review, with a non-significant increasing trend. The high CFR can be due to poor awareness of the disease (29,30), leading to delayed presentation, inadequate infection prevention and control (IPC) practices and poor health-seeking behavior among the population.

Bleeding and central nervous system (CNS) symptoms as predictors of mortality in a previous study (31) are in line with the findings of this study. While seizures were found to be significantly associated with mortality, as supported by Okokhere *et al* (32) the mechanism through which it leads to death, remains unclear (33). In addition to farming being associated with lower mortality(26) in Ebonyi State, this study finds that individuals aged over 44 years are more likely to die from LF.

### Study limitations and generalizability of findings

Although the findings provide valuable insights into the context of LF disease in Ebonyi State, they may not be generalizable across the country. The data analyzed was specific only to Ebonyi State and may not be representative outside the area of research due to differences in demography, environmental and cultural practices.

## Conclusion

LF in Ebonyi State peaks between November and March each year. LF incidence across the State was spatially heterogeneous, with consistently lower rates in southern LGAs and significant clustering of high-incidence LGAs in the central and northern regions. Interventions such as education and massive sensitization should focus on the productive age group, secondary school students, manual laborers, and high-burden LGAs that contribute to the high burden during peak periods. Training of healthcare workers in rural LGAs is advised to ensure early detection and referral of suspected cases. Further studies may be necessary to understand the absence of cases in Ivo LGA.

## Data Availability

The Lassa fever surveillance data that support the findings of this study are owned by the Nigeria Centre for Disease Control and Prevention and are not publicly available due to privacy and ethical restrictions. Data are available from NCDC through the Ebonyi State Ministry of Health upon reasonable request and with permission of the relevant data custodian.

## Notes

### Competing Interest Statement

The authors have declared no competing interest.

### Funding Statement

The authors received no specific funding for this work.

### Author Declarations

Ethics clearance was granted by the ITM IRB with No IRB/RR/AC/025/1749/24 and the Ebonyi State Health Research Ethics committee with protocol number EBSHREC/0061/052

